## Supplementary Tables and Figures for "Broad Clinical Manifestations of Polygenic Risk for Coronary Artery Disease in the Women’s Health Initiative"

[1]VA Palo Alto Health Care System, Palo Alto, CA
[2]Department of Medicine, Division of Cardiovascular Medicine, Stanford University School of Medicine, Stanford, CA
[3]Department of Epidemiology and Population Health, Stanford University School of Medicine, Stanford, CA
[4]Herbert Wertheim School of Public Health and Human Longevity Science, University of California, San Diego, La Jolla, CA
[5]Department of Epidemiology, Brown University, Providence, RI
[6]Fred Hutchinson Cancer Research Center, Seattle, WA
[7]Department of Medicine, Brigham and Women's Hospital, Harvard Medical School, Boston, MA
[8]Gladstone Institute, San Francisco, CA
[9]Department of Epidemiology and Biostatistics, University of California San Francisco, San Francisco, CA
* These authors contributed equally

**Supplementary Table 1. Overview of Genetic Studies Used to Define Study Population**

| (WHI Study #) | Hip Fracture GWAS | GARNET | WHIMS+ | GECCO | MOPMAP | Breast Cancer Post GWAS |
|---|---|---|---|---|---|---|
| | (BA3) | (M13) | (W63) | (AS224) | (AS264) | (M18) |
| **Study that funded GWAS** | WHI-BAA3 National Heart, Lung, and Blood Institute (NHLBI) | WHI-GARNET - National Human Genome Research Institute (NHGRI) | NHLBI | WHI-AS224 - National Cancer Institute (NCI) | WHI-AS264 - National Institute of Environmental Health Sciences (NIEHS) and Univ of North Carolina | M18 - NIH |
| **GWAS platform** | Illumina 550K and 610K | Illumina HumanOmni1-Quad v1-0 B | HumanOmniExpress Exome-8v1_B | Illumina 610 and Cytochip 370K | Affymetrix Gene Titan, Axiom Genome-Wide Human CEU I Array Plate | *llumina OncoChip* |
| **Design** | Case-control | Case-control (4 case groups) | Cohort | Case-control | Case-Control | Case-Control |
| **Phenotype for cases** | Hip Fracture | Type 2 Diabetes, Myocardial Infarction, Stroke, Venous Thrombosis | NA | Colorectal cancer | Ventricular Ectopy (ever) | Breast Cancer |
| **Other sample details** | NA | Hormone Therapy Clinical Trial | Hormone Therapy Clinical Trial | NA | Controls selected within centers, years, seasons and visit years in which cases originated | NA |
| **Self-identified race/ethnicity** | Mostly non-Hispanic white | Mostly non-Hispanic white | Non-Hispanic white | Non-Hispanic white | Non-Hispanic white | Non-Hispanic white |
| **Sample size** | 3,690 | 4,883 | 5,687 | 2,493 | 3,069 | 9,553 |

**Supplementary Table 2. Questions/forms used to identify subjects with a known or likely history of atherosclerotic cardiovascular disease (ASCVD) at baseline.**

| Source | Disease | Question |
|---|---|---|
| Form 2 - Eligibility Screening | Stroke ever | Did a doctor ever say that you had a stroke? |
| Form 2 - Eligibility Screening | Stroke last 6 months | Did you have a stroke in the last 6 months? |
| Form 2 - Eligibility Screening | TIA ever | Did a doctor ever say that you had a small stroke that lasted less than 24 hours? This is sometimes called a transient ischemic attack or TIA. |
| Form 2 - Eligibility Screening | TIA last 6 months | Did you have a TIA in the last 6 months? |
| Form 2 - Eligibility Screening | MI ever | Did a doctor ever say that you had a heart attack? This is sometimes called a coronary, myocardial infarction, or MI. |
| Form 2 - Eligibility Screening | MI last 6 months | Did you have a heart attack in the last 6 months? |
| Form 30 - Medical History | Cardiac arrest ever | Please mark the conditions or procedures below that a doctor said you had. Cardiac arrest (where your heart stopped and needed to be restarted) |
| Form 30 - Medical History | Coronary bypass surgery ever | Please mark the conditions or procedures below that a doctor said you had. Heart bypass operation or coronary bypass surgery for blocked or clogged arteries in you heart |
| Form 30 - Medical History | Angioplasty of coronary arteries ever | Please mark the conditions or procedures below that a doctor said you had. Angioplasty of the coronary arteries (opening the arteries of the heart with a balloon or other device, sometimes called a PTCA) |
| Form 30 - Medical History | Carotid endarterectomy/ angioplasty ever | Please mark the conditions or procedures below that a doctor said you had. Carotid endarterectomy or carotid angioplasty (operation for blockage or narrowing of the arteries in your neck) |
| Form 30 - Medical History | Aortic aneurysm ever | Please mark the conditions or procedures below that a doctor said you had. Aortic aneurysm |
| Form 30 - Medical History | Angina ever | Did a doctor ever say that you had angina (chest pains from a heart problem)? |
| Form 30 - Medical History | Pills for angina now | Do you now take pills for angina? |
| Form 30 - Medical History | Peripheral arterial disease ever | Did a doctor ever say that you had claudication or peripheral arterial disease (poor blood flow to the legs or blocked or narrowed arteries to the legs)? Do not include varicose veins or phlebitis. |
| Form 30 - Medical History | Angiography for PAD ever | For the above condition, have you ever had: Angiography (dye in the arteries of the legs)? |
| Form 30 - Medical History | Angioplasty for PAD ever | For the above condition, have you ever had: Angioplasty (balloon catheter to open blockage)? |
| Form 30 - Medical History | Surgery to improve flow to legs for PAD | For the above condition, have you ever had: Surgery to improve blood flow in your legs (do not include surgery for varicose veins)? |
| Form 30 - Medical History | CABG/PTCA Ever | Computed from Form 30, questions 3.1.4 and 3.1.5. Indicator for whether the participant has a history of either CABG or PTCA. |

TIA = transient ischemic attack; MI = myocardial infarction; PAD = peripheral arterial disease; CABG = coronary artery bypass; PTCA = Percutaneous Transluminal Coronary Angioplasty

**Supplementary Table 3. Cohort characteristics.** The study cohort was selected from the group of genotyped participants with inferred similar genetic ancestry. Non-genotyped participants are shown for comparison.

| **Characteristic** | Study Cohort, no ASCVD at enrollment (n = 21,863) | Genotyped participants with inferred similar genetic ancestry (N=24,693) | Non-genotyped participants, self-identified non-Hispanic white (N=109,037) |
|---|---|---|---|
| Age | 65.3 ± 6.3 | 65.66 ± 6.9 | 63.10 ± 7.2 |
| Self-identified non-Hispanic White | 21,698 (99.2) | 24,504 (99.2) | 109,037 (100) |
| Clinical Trial Status | 15,068 (68.9) | 16,959 (68.7) | 38,744 (35.5) |
| **Health traits** | | | |
| Total cholesterol | 223.4 ± 42.9 | 223.3 ± 43.0 | 221.3 ± 42.9 |
| LDL-C | 139.7 ± 39.0 | 139.7 ± 39.0 | 135.6 ± 39.6 |
| HDL-C | 55.0 ± 14.3 | 54.6 ± 14.3 | 56.3 ± 15.4 |
| Triglycerides | 143.0 ± 83.3 | 145.4 ± 84.2 | 136.4 ± 82.1 |
| Systolic Blood Pressure | 126.0 ± 6.0 | 129.2 ± 18.1 | 126.6 ± 17.8 |
| Body Mass Index | 28.5 ± 6.0 | 28.2 ± 5.8 | 27.5 ± 5.7 |
| **Parent study outcomes** | | | |
| Clinical MI | 1,309 (6.0) | 1,673(6.8) | 3,832 (3.5) |
| Breast Cancer | 4,928 (22.5) | 5,425 (22.0) | 6,241 (5.7) |
| Colon Cancer | 910 (4.20) | 1,010 (4.1) | 1,380 (1.3) |
| Diabetes | 2,168 (9.9) | 2,610 (10.6) | 10,035 (9.3) |
| Hip Fracture | 2,167 (9.9) | 2,575 (10.4) | 1,628 (1.5) |

ASCVD = atherosclerotic cardiovascular disease; LDL-C = low-density lipoprotein cholesterol; HDL-C = high-density lipoprotein cholesterol; MI = myocardial infarction

**Supplementary Table 4. Associations between polygenic risk for coronary artery disease and incident adjudicated cardiovascular diseases without risk factor adjustment.**

| Outcome | Cases | Controls | OR | LCI | UCI | P value |
|---|---|---|---|---|---|---|
| Coronary revascularization | 1,530 | 20,333 | 1.55 | 1.46 | 1.64 | 6.2E-53 |
| PTCA | 1,150 | 20,713 | 1.52 | 1.43 | 1.62 | 6.8E-39 |
| Myocardial infarction | 1,309 | 20,554 | 1.43 | 1.35 | 1.52 | 1.3E-31 |
| Coronary heart disease | 1,792 | 20,071 | 1.33 | 1.26 | 1.40 | 1.1E-26 |
| CABG | 501 | 21,362 | 1.56 | 1.42 | 1.71 | 1.8E-20 |
| All angina | 620 | 21,243 | 1.41 | 1.30 | 1.54 | 9.8E-16 |
| Ischemic stroke | 1,108 | 20,755 | 1.16 | 1.08 | 1.23 | 8.4E-06 |
| All stroke | 1,457 | 20,406 | 1.14 | 1.07 | 1.20 | 1.1E-05 |
| Carotid disease | 293 | 21,570 | 1.20 | 1.06 | 1.35 | 3.0E-03 |
| Peripheral artery disease | 275 | 21,588 | 1.21 | 1.06 | 1.37 | 3.2E-03 |
| Congestive heart failure | 579 | 21,284 | 1.09 | 1.00 | 1.19 | 6.0E-02 |
| TIA | 213 | 21,650 | 1.14 | 0.99 | 1.32 | 6.5E-02 |
| Angina without revascularization | 191 | 21,243 | 1.11 | 0.96 | 1.29 | 1.7E-01 |
| Valvular disease | 174 | 21,689 | 1.11 | 0.95 | 1.30 | 1.9E-01 |
| Deep vein thrombosis | 519 | 21,344 | 0.97 | 0.88 | 1.06 | 4.8E-01 |
| Hemorrhagic stroke | 214 | 21,649 | 0.97 | 0.84 | 1.11 | 6.3E-01 |
| Pulmonary embolus | 399 | 21,464 | 0.99 | 0.90 | 1.10 | 9.1E-01 |

**Supplementary Table 5. Sensitivity analysis of adjudicated cancer outcomes among the subset of women genotyped with the Oncochip.**

| Outcome | Cases | Controls | Odds ratio | LCI | UCI | P value |
|---|---|---|---|---|---|---|
| Any cancer | 4150 | 2808 | 0.96 | 0.91 | 1.01 | 0.1 |
| Breast cancer | 3734 | 3224 | 0.97 | 0.92 | 1.02 | 0.2 |
| Lung cancer | 167 | 6791 | 0.92 | 0.78 | 1.07 | 0.3 |

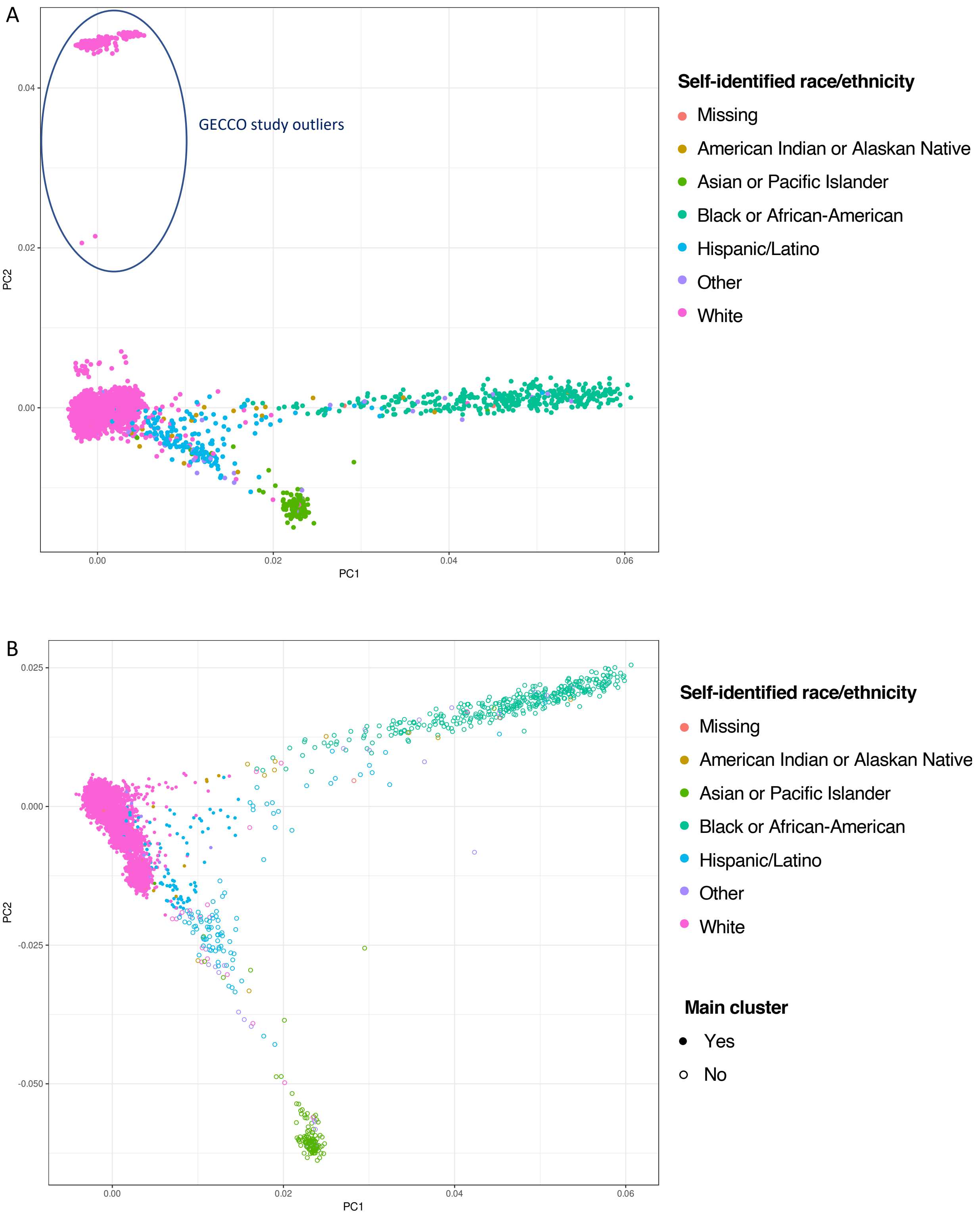


**Supplementary Figure 1. Principal components analysis of the genotyped cohort.** (A) In the initial analysis, 472 outliers from the GECCO study were identified. These subjects were removed. (B) Principal components were rerun. A cohort of 24,693 subjects (main cluster) were identified as having similar inferred genetic ancestry by the Mahalanobis distance method.

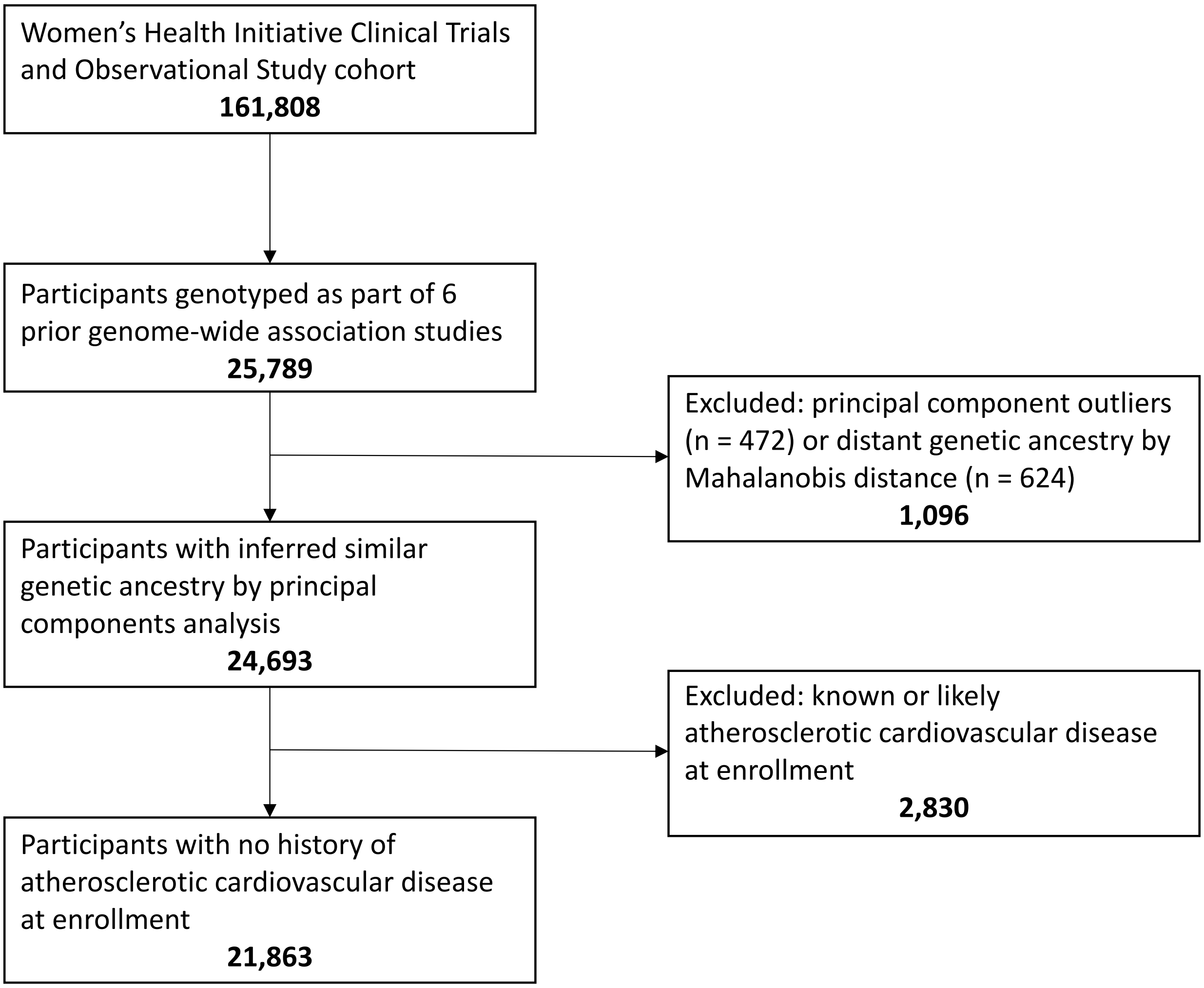


**Supplementary Figure 2. Flow diagram of Women's Health Initiative subjects selected for this study.**

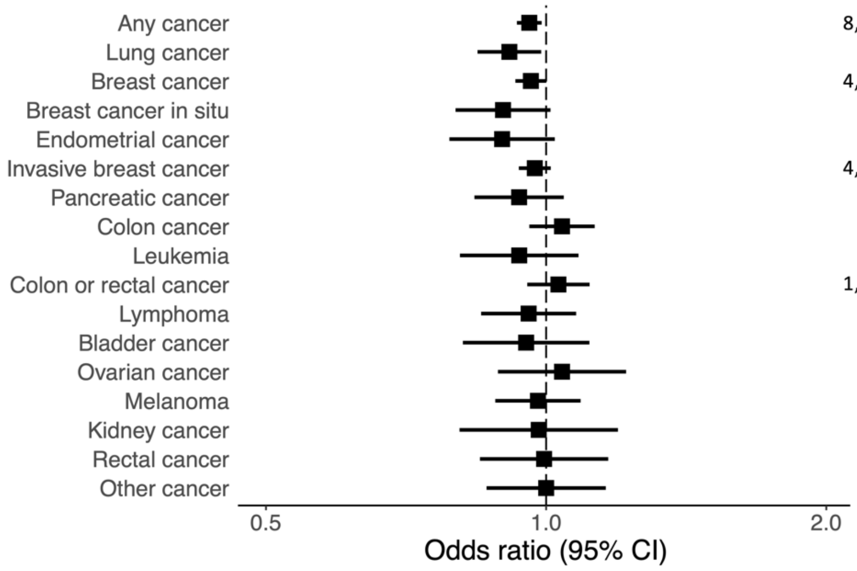


| | Cases | Controls | OR (95% CI) | P value |
|---|---|---|---|---|
| Any cancer | 8,294 | 13,569 | 0.96 (0.93-0.99) | 7.7E-03 |
| Lung cancer | 696 | 21,167 | 0.91 (0.84-0.99) | 2.4E-02 |
| Breast cancer | 4,928 | 16,935 | 0.96 (0.93-1.00) | 5.4E-02 |
| Breast cancer in situ | 309 | 21,554 | 0.90 (0.80-1.01) | 7.5E-02 |
| Endometrial cancer | 248 | 21,615 | 0.90 (0.79-1.02) | 0.10 |
| Invasive breast cancer | 4,686 | 17,177 | 0.97 (0.93-1.01) | 0.16 |
| Pancreatic cancer | 358 | 21,505 | 0.94 (0.84-1.04) | 0.24 |
| Colon cancer | 910 | 20,953 | 1.04 (0.96-1.13) | 0.34 |
| Leukemia | 195 | 21,668 | 0.94 (0.81-1.08) | 0.37 |
| Colon or rectal cancer | 1,151 | 20,712 | 1.03 (0.95-1.11) | 0.44 |
| Lymphoma | 307 | 21,556 | 0.96 (0.85-1.08) | 0.47 |
| Bladder cancer | 172 | 21,691 | 0.95 (0.81-1.11) | 0.54 |
| Ovarian cancer | 168 | 21,695 | 1.04 (0.89-1.22) | 0.63 |
| Melanoma | 383 | 21,480 | 0.98 (0.88-1.09) | 071 |
| Kidney cancer | 110 | 21,753 | 0.98 (0.81-1.19) | 0.85 |
| Rectal cancer | 193 | 21,670 | 0.99 (0.85-1.17) | 0.95 |
| Other cancer | 194 | 21,669 | 1.00 (0.86-1.16) | 1.00 |

**Supplementary Figure 3. Associations between polygenic risk for coronary artery disease and incident adjudicated cancers in the Women's Health Initiative.** Outcomes with at least 100 incident cases were considered. Odds ratios are per 1 standard deviation increase in polygenic risk score.
